# Adolescents’ Use of Music for Pain Management

**DOI:** 10.1101/2025.02.04.25321472

**Authors:** Rebecca J. Lepping, Lora L. Black, Kymberly A. Kline, Deanna Hanson-Abromeit, Andrea L. Chadwick, Dustin P. Wallace, William R. Black

## Abstract

To investigate the experiences of adolescents with chronic pain who participated in an intensive interdisciplinary pain treatment program, this secondary study analyzes the themes that emerged regarding the spontaneous utilization of music in coping strategies for chronic pain. During research interviews focused on coping skills and treatment engagement, participants spontaneously reported using music as an effective coping strategy for managing pain. A deductive thematic analysis revealed key themes related to their usage, including using music as a distractor, motivator and in other ways as coping strategies. Since participants indicated that music is essential to their experiences of coping with pain, incorporating these strategies could improve the effectiveness of treatment protocols. To this end, further investigation is necessary to assess the impact of music on adolescents with chronic pain, focusing on its role in enhancing interdisciplinary treatment.

---

Chronic pain, defined as pain that occurs for at least three months, impacts up to 15-44% of adolescents aged 10-18.^1,2^ In this age range, the occurrence of chronic pain is associated with challenges in academic performance, increased school absences, and psychosocial difficulties including anxiety, depression, sleep concerns, and struggles with peer relationships.^3-8^ Adolescents with chronic pain are also at an increased risk for having chronic pain as an adult which can lead to additional healthcare costs and lower educational and/or vocational attainment as they age. As well, it is estimated that pediatric chronic pain costs approximately $19.5 billion annually in the United States.^9^ Thus, it is important that pediatric patients with chronic pain have access to appropriate treatment to help minimize the impact that these conditions have on their life and to promote optimal functioning.

For young people who are highly impaired by chronic pain, treatment may include Intensive Interdisciplinary Pain Treatment (IIPT).^10^ IIPT programs typically include physical activity, desensitization, psychotherapy, education about pain mechanisms, learning and practicing stress management techniques, and an overall focus on moderation and normal functioning even when experiencing pain. As part of IIPT, patients are encouraged to develop and regularly use psychological, relaxation-based, and expressive coping skills to aid in the management of their conditions. These programs teach a variety of skills, allowing patients to identify which are most appealing and effective, increasing the likelihood of incorporating them into their daily lives after graduating from IIPT.

Music may be a particularly effective coping skill for adolescents with chronic pain.^11^ Music therapy has been utilized in multiple pediatric populations^12-14^ and is associated with improved disease coping during their treatment,^12,14^ and overall quality of life.^11^ Music and music therapy programs may also be effective for pain coping in adolescent chronic pain and are included in some IIPTs. However, there is limited information about how children and adolescents with chronic pain use music outside of formal treatment programs.

Analgesia from music-based interventions (MBIs) may be due to psychological or physiological effects.^15^ Contextually, music unfolds over time in predictable ways leading to expectancies and providing context for what is coming next. This effect is amplified when the music is familiar, as it can evoke additional resources such as future thinking or enhancing sentimental feelings. Music listeners experience enhanced analgesic effects when they select their own music. Cognitive factors are also at play. Music can act as a cognitive distraction, redirecting attention away from painful feelings. This phenomenon is not exclusive to music; similar effects can be achieved through other distraction methods, such as reading or listening to nature sounds.^16^ Emotion is a key component of the music experience as music has a compelling ability to trigger emotional responses.^17,18^ For instance, emotionally positive music that is liked by the listener and has low arousal levels produces the strongest analgesic effects.^19^ This has also been demonstrated to have clinical relevance as music listening interventions and music therapy have demonstrated effectiveness in alleviating anxiety and depression.^20,21^ Physiologically, the impact of music for pain may potentially be mediated through action on the parasympathetic nervous system, via vagal activation and lowering cardiac and respiration rates.^22^ There have also been physiological effects observed in the brain, with music linked to the release of endogenous opioids and dopamine and activation of the descending pain modulatory system during the experience of pain.^23,24^

Given the multiple components of IIPT and the various strategies that are provided as part of these program, we were interested in understanding how much adolescents who complete these programs go on to use music. To that end, we conducted a secondary analysis of interviews of adolescents who had completed a program to specifically examine uses of music in their responses to a general query about strategies that they are using to support their functional and health-related goals.

## Methods

The parent study interviewed a group of adolescents with Chronic Musculoskeletal Pain (CMSKP) who had completed an IIPT program at a large children’s hospital. The IIPT program provides 8 hours of treatment, 5 days a week, with treatment duration ranging from 4-6 weeks depending on each patient’s progress with specific functional goals. Treatment includes 4-5 hours per day of physical and occupational therapy with a focus on exercise, desensitization, and a focus on normalizing physical functioning that had been impaired by pain. The other 3-4 hours of each day includes individual and group psychotherapy (5 hours per week), guided relaxation and meditation (2 hours per week), yoga (2 hours per week), art therapy (2 hours per week) and music therapy (2 hours per week), all provided by licensed therapists. The program does not specifically focus on pain management, but instead focuses on improved functioning, returning to valued activities, and gaining confidence to adhere to recommendations following the program which include daily exercise, desensitization, and proactive use of stress management strategies.

While analyzing the interview data for themes focused on the use of coping skills and engagement in recommended treatments for chronic pain, themes emerged regarding participants’ discussion of the role that music plays in their experience of chronic pain. Although participation in music therapy, art therapy, and other relaxation techniques (yoga, meditation, etc.) were part of the structured program, the interview question did not specifically ask about music. Participants’ self-reports identified unsolicited thematic usage as to how they incorporated music in an unstructured manner. These were unguided ways in which participants utilized music and unsolicited self-report of music usage incorporated into their experiences.

## Participants

Eligible participants were adolescents (ages 13-17 years) with a history of CMSKP who had completed an IIPT at the study’s institution and who had agreed to be contacted via email or recruitment flyers for future research. Ten participants enrolled in the study and completed the semi-structured interviews (*M*age = 16.45 years, *SD* = 1.35, range = 13.44 – 17.73). Ninety percent (*n* = 9) of the sample identified as female.

## Procedures

Study procedures occurred remotely. Participants were recruited via email (i.e., existing patients that had agreed to be contacted for future research participation) or were referred by providers in pain management clinics. After IRB-approved virtual consent, participants completed surveys assessing background information and provided additional information about the purpose and structure of the interview. Individual semi-structured interviews were conducted via an online HIPPA-compliant video communication platform (Microsoft Teams). Participants were informed that they could decline to answer any interview question or terminate the interview at any time. All study procedures were approved by the institution’s IRB.

## Semi-Structured Interviews

A trained psychology interviewer (JC) supervised by a licensed psychologist (WB) conducted the individual interviews, which ranged from 45-60 minutes in duration and were audio recorded for transcription. The interviewer followed a semi-structured interview script constructed to elicit participants’ lived experiences with CMSKP and their engagement in recommended treatments, including use of helpful pain-focused management and coping skills.

## Qualitative Data Analysis

While the original study was focused on the use of coping skills and engagement in recommended treatments for chronic pain and not on the use of music specifically, initial analysis indicated that 7 out of the 10 participants discussed the role of music in their experience of chronic pain without prompting. Thus, we conducted a secondary thematic analysis focused on participants’ experiences with music, art, and relaxation techniques in relation to their chronic pain without a predetermined framework. To systematically examine the use of music reported by these participants, we conducted a qualitative thematic analysis of the interviews specifically for the use of music, the functions of music in that moment, and the situational appropriateness of that as a strategy. As music therapy and art therapy were provided in equal proportion during the program (1-2 hours per week), we also examined the interview responses for uses of art.

Interview recordings were transcribed verbatim by a third-party transcription service and then checked for accuracy (JC). Identifying information was removed prior to transcript review and coding. Interview transcripts were analyzed in Dedoose (version 9.0.17, Los Angeles, CA: SocioCultural Research Consultants, LLC) using deductive thematic analysis to identify, analyze, and report themes found within the data from a predefined theoretical framework.^25^ For the parent study, we analyzed the transcripts from a pain resilience framework, highlighting various psychological factors that enable continued engagement in activities and behaviors while a person is experiencing pain.^26,27^ Two team members (JC and LB) reviewed the transcripts and developed the initial codes, which were discussed and agreed upon by team members. All initial codes that discussed music were then analyzed separately by two team members (LB and WB) to be independently categorized into music-focused codes. Any discrepancies in coding were discussed until consensus was reached. In the event consensus was not reached, a third team member (RL) reviewed the responses and codes to decide on final coding categorization. All themes were developed from the iterative review of the seven participants who discussed music during their interview, where coders grouped similar responses together into codes which were then assigned a theme to best represent the group of responses.

## Results

### Qualitative themes

#### 1. Using music as a distractor or to help pass the time

Some of the participants reported that they used music to help distract them from the pain or when they were in overstimulating situations. Similarly, other participants described using music to help pass the time when engaging in other management techniques for their chronic pain.

> *“But like if I’m in public of course, I’m usually -- I’m just really big on distracters*. ***Music***, *anything helps me focus on things…” (Participant 2)*
>
> *“I … listen to* ***music*** *while going on runs. It just passes time.” (Participant 9)*
>
> *“Doing something enjoyable while also doing something not so enjoyable. It makes it a little easier. So that would be one thing I’d recommend is just having something to do. So listening to* ***music*** *… having something else going on where it’s like I’m having some -- this isn’t the enjoyment that I want, but it’s enjoyment while I’m doing something I need to do.” (Participant 3)*

#### 2. Using music as a motivator

Participants also discussed how they would listen to music to help motivate them to keep using pain management recommendations, especially difficult ones such as physical activity. Of note, some also discussed the impact that music can have on both their mood and motivation.

> *“So even when walking alone, I have a* ***playlist*** *that’s just full of super upbeat, cheery* ***music*** *with a beat that will just keep me going the entire time. I definitely don’t play slow* ***songs*** *or anything that you just*… *But just having that good* ***playlist***.*” (Participant 2)*
>
> *“I remember sometimes you guys would let us listen to* ***music*** *while we biked or elliptical or whatever that was, naturally boosted me. I remember that, like, I listened to some* ***motivational music***, *for me, it was Fight Song by Rachel Platten, that was my fight song, like, I’m going to get through this, this is my fight song. Yeah, that was my fight song. And so, I was able to… go ahead and listen to some music, and that truly boosted my mood in order to get the work done.” (Participant 6)*
>
> *“I like [engaging in physical activity] by myself and blasting* ***music***. *I find is what really just gets me on the zone.” (Participant 7)*

#### 3. Using music in other ways as a coping mechanism

Other participants discussed how they incorporated music into their repertoire of coping mechanisms aside from just listening to it, such as analyzing lyrics or playing an instrument.

> *“So a few days after I graduated from the program I experienced a really bad pain flaring…. And I was super upset about it… but with some coping strategies I’ve learned from the [IIPT] program, I picked myself up, stop feeling sorry for myself, said, okay, they gave me these resources need to start using them. … one of the things I loved was with [the music therapist] when we* ***listen to music*** *and …we just looked into the* ***lyrics***, *it’s a whole new meaning.” (Participant 7)*
>
> *“we decided I just could not do yoga or meditate, we decided that was just not in the picture. Yeah. And so, we had to come up with different ways that I could do that were reasonable. So, I’d listen to* ***music*** *or* ***play the piano*** *or write.” (Participant 1)*

#### 4. Art for relaxation, distraction, or in combination with music

Four participants who had described their use of music also described using art, including drawing, painting, or needlework alone or in combination with music as coping mechanisms.

> *“I really don’t know because I didn’t do physical activity, you know just by myself because I didn’t do anything. I’d rather sit at home and* ***draw*** *or something.” (Participant 1)*
>
> *“wool needle felting [to help me] settle with the pain” (Participant 3)*
>
> *“the one I use all the time is* ***music or drawing***.*” “Just distracting my mind is what helps me the most. So, if I just think about something else or really focus on something else, like, for example* ***art***, *yeah, it really helps.”(Participant 5)*
>
> *“I think one of my main coping mechanisms before I came to your program and you gave me much better ones was definitely a lot of reading, a lot of sitting down, a lot of -- I even got used to* ***painting and drawing*** *at that time. Like, you know, I never did that before, but when I was doing that, when I was, like, not even being able to move, I think like, just like, you know, either reading, drawing, stuff like that.” (Participant 6)*

## Discussion

During a semi-structured follow-up interview focusing on participants’ experiences with CMSKP, participants who had previously completed an IIPT program mentioned music as a resource for managing pain. These were spontaneous responses about music and art to a general question about coping strategies for pain, and while music therapy was a component of the treatment program, it was not specifically described during the program as being an effective strategy for managing pain. Participants discussed the role of music in their experience with chronic pain without instruction on how to implement it as a strategy. Themes that emerged from the participants use of music varied including distraction, motivation, coping mechanisms and psychological function of the music.

For participants who mentioned using music to distract themselves from the pain or to reduce social stress in social situations, treatment often included a combination of active and passive usage of coping strategies. For example, studies have shown that for those engaged in recommended exercise strategies, music can be used as distraction from the physical symptoms they are having, while coping to help them with the emotional part of the pain experience.^28^ In this way, listening to music can be a perceived way to control the external situation by managing inner/outer spaces, avoiding stressors from the environment, and self-regulating. Additionally, it is not always feasible to engage in relaxation strategies like yoga or deep breathing, so having a mix of skills or tools available that can best fit the situation can be helpful. Other participants used music as a distraction from the passage of time during over stimulating environments. It has been shown that when individuals feel they have little control over their environment, music becomes an important tool for managing their internal mood to cope with the lack of control from the external world.^28^ In this way, music may function as a mindfulness technique to bring engagement into the present moment and away from the focus on pain.

Participants also highlighted how they use music to motivate themselves to “push past the pain.” Music appears to be especially valuable for individuals who are either developing their sense of agency, such as youth, or facing a decrease in their ability to control actions and environment due to illness or difficult personal circumstances.^29^ By harnessing its emotional power, music can motivate individuals into actions.^30^ For some, solitary listening can be considered a significant aspect of youth development, as the solitary experience fosters independence, serves as self-discovery, and personal mood regulation.^31^ Creating personalized playlists further individualizes the experience of music listening by enhancing the effectiveness of a coping strategy, for example, creating a sad song play list to intentionally reflect mood and can be used as a coping strategy when they are sad.

One such need, accessibility and societal approval, along with the cultural phenomenon that teens use music to form identity, voice protest or express thoughts that may conflict with societal norms, which can be accessed through listening, composing and performing make music unique in its usability compared to other coping methods. Additionally, portability makes it easier to incorporate music into daily routines as a coping mechanism. Incorporating music into the routine can integrate this mechanism into life.

## Limitations

There are some limitations to consider while evaluating these results. First, the relatively small sample size was appropriate for exploring themes but would not allow for generalizability of the findings. Collecting a larger sample size may generate findings that more broadly reflect how adolescents with CMSKP are using music for coping with pain. Additionally, participants were not specifically asked about their music usage; therefore, the data collected only reflects instances where adolescents with CMSKP mentioned music spontaneously. This may not fully capture the extent to which they engage with music in their lives as a coping strategy.

## Conclusion

The present investigation is suggestive that more research is needed to investigate the role of music in the lives of adolescents with CMSKP. By further exploring how music can make other parts of the interdisciplinary treatment more tolerable or effective. Additionally, by including music into treatment and management plans for distraction, motivation, and coping strategies. Future studies should delve deeper into the topic, potentially by utilizing another qualitative approach to gain smore comprehensive insights into the specific aspects or characteristics of music that provide the most benefit. Understanding the underlying physical and neurological mechanisms of music and art for pain management is key for incorporating these tools into prescriptive and personalized medicine, as we may be able to better understand unique, person specific responses to music and then teach the use of music as a coping strategy in alignment with their preferences. Therefore, future research using functional magnetic resonance imaging (fMRI) or electroencephalography (EEG) to understand how music changes the brain response to pain would be warranted.

## Data Availability

All data produced in the present study are available upon reasonable request to the corresponding authors

## Acknowledgements

The authors thank Ashley Scheufler, MT and Jennifer Christofferson, PhD for their contributions to this project. Research reported in this publication was supported with funding from the University of Kansas Medical Center, Department of Pediatrics’ Pediatric Research Grant and the National Center for Advancing Translational Sciences of the National Institutes of Health under the Award Number UL1TR002366 to Rebecca Lepping. The content is solely the responsibility of the authors and does not necessarily represent the official views of the National Institutes of Health, Children’s Mercy Kansas City, or the University of Kansas Medical Center. We gratefully acknowledge the participants and their families for sharing their stories and experiences.

## Notes

### Competing Interest Statement

The authors have declared no competing interest.

### Author Declarations

Ethics committee/IRB of Children's Mercy Hospital Kansas City gave ethical approval for this work.

## REFERENCES

1. Gobina I, Villberg J, Valimaa R, et al. Prevalence of self-reported chronic pain among adolescents: Evidence from 42 countries and regions. Eur J Pain. Feb 2019;23(2):316–326. doi:10.1002/ejp.1306

2. de la Vega R, Groenewald C, Bromberg MH, Beals-Erickson SE, Palermo TM. Chronic pain prevalence and associated factors in adolescents with and without physical disabilities. Dev Med Child Neurol. Jun 2018;60(6):596–601. doi:10.1111/dmcn.13705

3. Simons LE, Logan DE, Chastain L, Stein M. The relation of social functioning to school impairment among adolescents with chronic pain. Clin J Pain. Jan 2010;26(1):16–22. doi:10.1097/AJP.0b013e3181b511c2

4. Vervoort T, Logan DE, Goubert L, De Clercq B, Hublet A. Severity of pediatric pain in relation to school-related functioning and teacher support: an epidemiological study among school-aged children and adolescents. Pain. Jun 2014;155(6):1118–1127. doi:10.1016/j.pain.2014.02.021

5. Richardson PA, Birnie KA, Harrison LE, Rajagopalan A, Bhandari RP. Profiling Modifiable Psychosocial Factors Among Children With Chronic Pain: A Person-Centered Methodology. J Pain. Mar-Apr 2020;21(3-4):467–476. doi:10.1016/j.jpain.2019.08.015

6. Murray CB, Groenewald CB, de la Vega R, Palermo TM. Long-term impact of adolescent chronic pain on young adult educational, vocational, and social outcomes. Pain. Feb 2020;161(2):439–445. doi:10.1097/j.pain.0000000000001732

7. Logan DE, Simons LE, Stein MJ, Chastain L. School impairment in adolescents with chronic pain. J Pain. May 2008;9(5):407–16. doi:10.1016/j.jpain.2007.12.00318

8. Hoffart CM, Wallace DP. Amplified pain syndromes in children: treatment and new insights into disease pathogenesis. Curr Opin Rheumatol. Sep 2014;26(5):592–603. doi:10.1097/BOR.0000000000000097

9. Groenewald CB, Essner BS, Wright D, Fesinmeyer MD, Palermo TM. The economic costs of chronic pain among a cohort of treatment-seeking adolescents in the United States. J Pain. Sep 2014;15(9):925–33. doi:10.1016/j.jpain.2014.06.002

10. Simons LE. Growing up in the society of pediatric psychology: reflections of an early career psychologist. J Pediatr Psychol. Mar 2013;38(2):132–4. doi:10.1093/jpepsy/jss121

11. Scheufler A, Wallace DP, Fox E. Comparing Three Music Therapy Interventions for Anxiety and Relaxation in Youth With Amplified Pain. J Music Ther. Jun 14 2021;58(2):177–200. doi:10.1093/jmt/thaa021

12. Colwell CM, Edwards R, Hernandez E, Brees K. Impact of music therapy interventions (listening, composition, Orff-based) on the physiological and psychosocial behaviors of hospitalized children: a feasibility study. J Pediatr Nurs. May-Jun 2013;28(3):249–57. doi:10.1016/j.pedn.2012.08.008

13. Millett CR, Gooding LF. Comparing Active and Passive Distraction-Based Music Therapy Interventions on Preoperative Anxiety in Pediatric Patients and Their Caregivers. J Music Ther. Jan 13 2018;54(4):460–478. doi:10.1093/jmt/thx014

14. Whitehead-Pleaux AM, Baryza MJ, Sheridan RL. The effects of music therapy on pediatric patients’ pain and anxiety during donor site dressing change. J Music Ther. Summer 2006;43(2):136–53. doi:10.1093/jmt/43.2.136

15. Lunde SJ, Vuust P, Garza-Villarreal EA, Vase L. Music-induced analgesia: how does music relieve pain? Pain. May 2019;160(5):989–993. doi:10.1097/j.pain.000000000000145218

16. Villarreal EA, Brattico E, Vase L, Ostergaard L, Vuust P. Superior analgesic effect of an active distraction versus pleasant unfamiliar sounds and music: the influence of emotion and cognitive style. Plos One. 2012;7(1):e29397. doi:10.1371/journal.pone.0029397

17. Juslin PN, Sloboda JA. Music and emotion : theory and research. Series in affective science. Oxford University Press; 2001:viii, 487 p.

18. Reybrouck M, Eerola T. Music and Its Inductive Power: A Psychobiological and Evolutionary Approach to Musical Emotions. Front Psychol. 2017;8:494. doi:10.3389/fpsyg.2017.00494

19. Basinski K, Zdun-Ryzewska A, Majkowicz M. The Role of Musical Attributes in Music-Induced Analgesia: A Preliminary Brief Report. Front Psychol. 2018;9:1761. doi:10.3389/fpsyg.2018.01761

20. Aselton P. Sources of stress and coping in American college students who have been diagnosed with depression. J Child Adolesc Psychiatr Nurs. Aug 2012;25(3):119–23. doi:10.1111/j.1744-6171.2012.00341.x

21. Brandes V, Terris DD, Fischer C, et al. Receptive music therapy for the treatment of depression: a proof-of-concept study and prospective controlled clinical trial of efficacy. Psychother Psychosom. 2010;79(5):321–2. doi:10.1159/000319529

22. Ribeiro MKA, Alcantara-Silva TRM, Oliveira JCM, et al. Music therapy intervention in cardiac autonomic modulation, anxiety, and depression in mothers of preterms: randomized controlled trial. BMC Psychol. Dec 13 2018;6(1):57. doi:10.1186/s40359-018-0271-y

23. Blood AJ, Zatorre RJ. Intensely pleasurable responses to music correlate with activity in brain regions implicated in reward and emotion. Proc Natl Acad Sci U S A. Sep 25 2001;98(20):11818–23. doi:10.1073/pnas.19135589898/20/11818 [pii]

24. Salimpoor VN, Benovoy M, Larcher K, Dagher A, Zatorre RJ. Anatomically distinct dopamine release during anticipation and experience of peak emotion to music. Nat Neurosci. Feb 2011;14(2):257–62. doi:10.1038/nn.2726

25. Braun V, Clarke V. Using Thematic Analysis in Psycholgoy. Qualitative research in psychology. 2006;3(2):77–101.

26. Goubert L, Trompetter H. Towards a science and practice of resilience in the face of pain. Eur J Pain. Sep 2017;21(8):1301–1315. doi:10.1002/ejp.1062

27. Parsons RD, McParland JL, Halligan SL, Goubert L, Jordan A. The perception, understanding and experience of flourishing in young people living with chronic pain: A Q-methodology study. J Health Psychol. Oct 2024;29(12):1350–1364. doi:10.1177/13591053241237341

28. Saarikallio SH, Randall WM, Baltazar M. Music Listening for Supporting Adolescents’ Sense of Agency in Daily Life. Front Psychol. 2019;10:2911. doi:10.3389/fpsyg.2019.02911

29. Magee WL. Music-making in therapeutic contexts: Reframing identity following disruptions to health. In: MacDonald RAR, Hargreaves DJ, Miell D, eds. Handbook of musical identities. First edition. ed. Oxford University Press; 2017:624–641:chap 34.

30. Krueger J. Affordances and the musically extended mind. Front Psychol. Jan 6 2014;4:1003. doi:10.3389/fpsyg.2013.01003

31. Larson R. Secrets in the Bedroom - Adolescents Private Use of Media. J Youth Adolescence. Oct 1995;24(5):535-550. doi:10.1007/Bf01537055

